# Infection rate and Immune Response to SARS-CoV-2 in Canadian Retail Workers -Cohort Description

**DOI:** 10.1101/2023.08.18.23294172

**Authors:** Mathieu Thériault, Kim Santerre, Nicholas Brousseau, Samuel Rochette, Rabeea F. Omar, Joelle N. Pelletier, Caroline Gilbert, Jean-François Masson, Mariana Baz, Denis Boudreau, Sylvie Trottier

## Abstract

Retail workers are an understudied occupational group that may have been at increased risk of contracting SARS-CoV-2 during the COVID-19 pandemic. To explore this question, we set up a longitudinal cohort of participants working in this sector to document the rate of SARS-CoV-2 infection and the immune response to infection and/or vaccination. A total of 304 participants were recruited between April 20^th^, 2021 and October 22^nd^, 2021. They were invited to attend three visits (each separated by ∼12 weeks) during which they provided blood samples. Information was collected on participant characteristics, SARS-CoV-2 detection tests done, COVID-19 symptoms, and vaccination. An extension phase of two additional visits was carried out between March 15^th^, 2022 and October 3^rd^, 2022 to document the impact of the Omicron variant among the 198 participants who were still eligible for recruitment. Participants were aged 18 to 75 and worked in grocery stores, hardware stores, bars or restaurants within the Québec City metropolitan area (Canada). *Findings to date:* This article describes participants’ demographic, socioeconomic, behavioral, occupational and clinical characteristics, and their COVID-19 symptoms (when applicable), as well as SARS-CoV-2 vaccination status and any SARS-CoV-2 positive diagnostic test (i.e., PCR or rapid antigen) from the beginning of the pandemic until the last visit. *The goals of this study were:* to assess the rate of SARS-CoV-2 infection, and the immune response (innate and acquired) to SARS-CoV-2 infection or vaccination using a variety of techniques. The humoral immune response was measured by reference and experimental enzyme-linked immunosorbent assays, as well as microneutralization assays with live viruses and experimental pseudoneutralization with an angiotensin-converting enzyme 2-spike assay. Cellular immune response was evaluated by using the viral analog R848 to activate peripheral blood mononuclear cells and neutrophils.

## INTRODUCTION

During the SARS-CoV-2 pandemic, workers with client-facing duties were considered to be at greater risk of infection than those who worked remotely (1–3). Although most studies have focused on healthcare workers (HCW) (4, 5) many non-HCWs were also considered to be at risk due to their occupational exposure (6, 7).

Workers in the food and retail industry are an understudied occupational group that may have been at greater risk of contracting SARS-CoV-2. These workers often have below-average incomes, face precarious employment conditions and lack benefit packages to cover health-related absenteeism. At the beginning of the pandemic, these workers often had insufficient training and access to protective equipment to reduce exposure, compared to HCWs (8). However, risk is likely to vary from one sector to another. For example, grocery and hardware stores were considered essential services and therefore remained open throughout the pandemic, with public health measures (e.g., mask wearing) being enforced and generally well respected. In contrast, restaurants and bars were intermittently opened and closed by health authorities over the same period, and public health measures were more difficult to enforce due to the intrinsically social nature of these businesses and their main purpose — the consumption of food and drink — that precluded continuous mask wearing.

Early evidence confirms that the risk of occupational exposure was high for these workers. In a serological survey conducted in New York City prior to the approval of the first COVID-19 vaccine, the seroprevalence of anti-spike antibodies was higher among grocery store and restaurant workers than in most subgroups of HCWs (1). In another serosurvey conducted in Switzerland, kitchen staff and grocery store workers exhibited an above average seroprevalence compared to other essential workers (9). In the Netherlands, individuals working in the hospitality sector were more likely to have a positive PCR test result than those working in non-close-contact occupations (10). In Japan, restaurants and bars were the second most common setting of COVID-19 outbreaks after healthcare facilities (7, 11).

To date, no thorough investigation of SARS-CoV-2 exposure has been conducted among Canadian workers in grocery stores, hardware stores, bars or restaurants (12, 13). Accordingly, we set up a longitudinal cohort that investigated the rate of COVID-19 during the first years of the pandemic and the humoral and cellular immunity (innate and acquired) to SARS-CoV-2 in these workers. This article describes the experimental design of the project and the cohort of participants.

## Study design & method

### Participants and setting

Eligibility criteria included the following: (1) providing informed consent; (2) age ≥18 years; (3) working either on a full-time or part-time basis in a grocery store, hardware store, bar or restaurant located in the administrative regions of Capitale-Nationale and Chaudière-Appalaches that include and surround Québec City, Canada; (4) having a public-facing role in daily work-related activities; (5) having worked ≥20 days between February 1^st^, 2020 and the first visit; and (6) having no history of hospitalization due to COVID-19.

Participants were recruited using a variety of strategies: 1) an online recruitment campaign conducted by a student-run communication agency; 2) email invitations to members of partner union organizations - *Confédération des syndicats nationaux* (CSN) and to sectoral organizations of hardware store workers - *Association Québécoise de la quincaillerie et des matériaux de construction* (AQMAT); and 3) email information to all students and employees at *Université Laval* and at the *Centre Hospitalier Universitaire de Québ*ec in order to publicize the study.

### Design and procedures

The study was initially designed as a prospective cohort study with three sampling visits, each separated by 12±2 weeks. In response to the emergence of Omicron, an extension of two visits was proposed to the participants who were still eligible for recruitment. An additional COVID-19 visit (VCoV) was also planned shortly after the occurrence of any SARS-CoV-2 infection during the study period. The study enrolled between April 20^th^, 2021 and October 22^nd^, 2021. The first three visits spanned from April 20^th^, 2021 to May 9^th^, 2022. The extension phase was carried out from March 15^th^, 2022 to October 3^rd^, 2022 (Fig 1).

**Figure 1.**
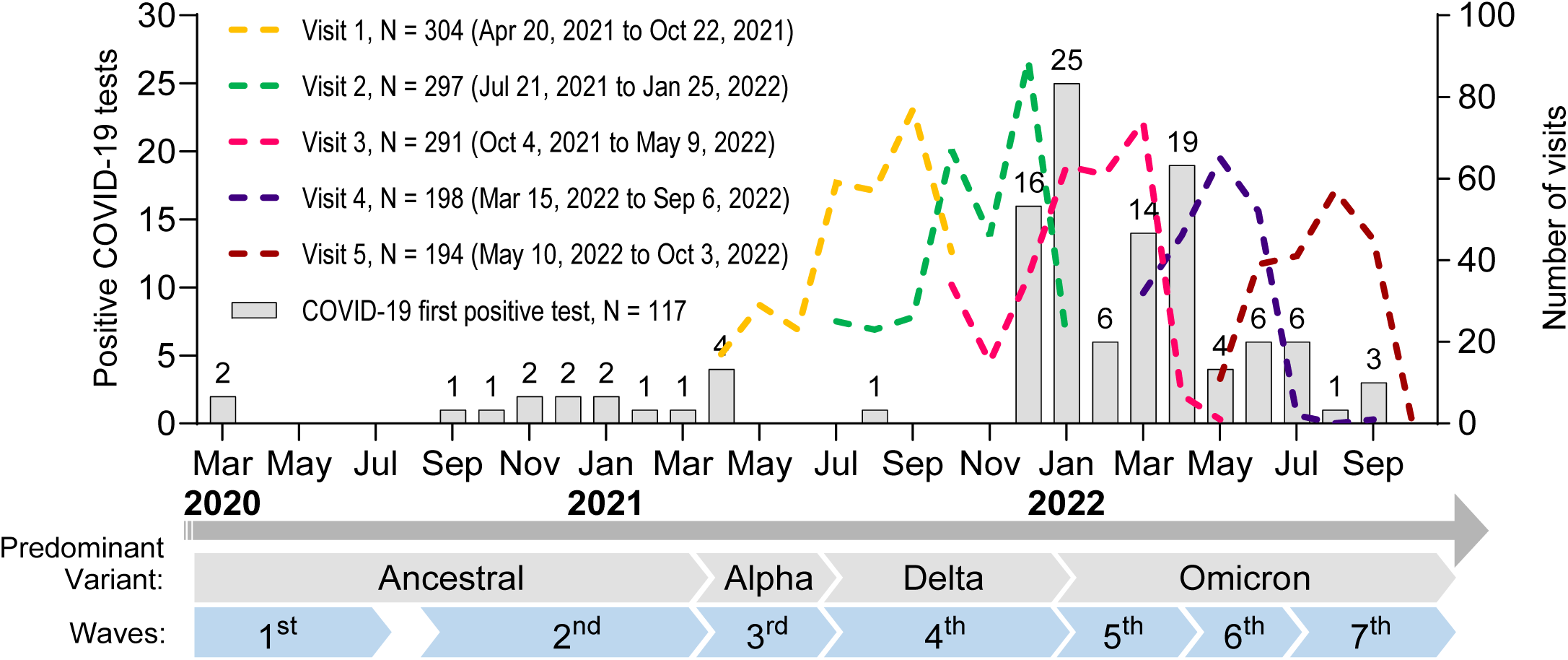
Timeline of the study illustrating the visits (colored dash lines) and COVID-19-positive tests (grey bars)

At the first visit (“V1”), participants signed an informed consent form and were then interviewed by trained nurses to obtain information on demographic, socioeconomic, behavioral, occupational and clinical variables (Table 1). The questionnaires were adapted from those suggested by our funding body, the COVID Immunity Task Force (CITF) (14). At or after the third visit (“V3”), eligible participants received information about the extension of the study and signed a new informed consent if they were interested in participating.

**Table 1.** Participant visits.

|  | Visit 1<br>N=304 | Visit 2<br>N=297 | Visit 3<br>N=291 | Visit 4<br>N=198 | Visit 5<br>N=194 | COVID-19<br>visit (VCoV)<br>N=15 |
| --- | --- | --- | --- | --- | --- | --- |
| **Retention** | 100% | 98% | 96% | 100% | 98% | - |
| **Eligibility assessment** | X |  |  | X |  |  |
| **Consent form** | X |  |  | X |  |  |
| **Participant characteristics** |  |  |  |  |  |  |
| Demographic data | X |  |  |  |  |  |
| Household data | X |  |  |  |  |  |
| Clinical data | X |  |  |  |  |  |
| Occupational data | X | X | X | X | X | X |
| **Retrospective questionnaire form** |  |  |  |  |  |  |
| COVID-19 symptoms | X | X | X | X | X | X |
| Positive SARS-CoV-2 detection tests | X | X | X | X | X | X |
| SARS-CoV-2 vaccine | X | X | X | X | X | X |
| **Cross-section interventions** |  |  |  |  |  |  |
| Humoral immunity blood samples | X | X | X | X | X | X |
| Cellular immunity blood samples | X |  | X |  | X | X |
| SARS-CoV-2 PCR test at visit |  |  |  | X | X |  |

At the second (“V2”), third (“V3”), fourth (“V4”) and fifth visits (“V5”), participants completed an abridged version of the V1 questionnaire that focused on SARS-CoV-2 vaccines received, COVID-19 symptoms, SARS-CoV-2 detection (PCR or rapid antigen) tests done, exposure and associated risk factors (Table 1). Blood was drawn to study humoral immunity (i.e., at V1 to V5) and cellular immunity (i.e., at V1, V3, and V5) to SARS-CoV-2. Additional PCR tests were carried out at V4 and V5 to detect asymptomatic carriers.

COVID-19 visit (VCoV), were scheduled for participants developing a SARS-CoV-2 infection during the study, and took place at a median time of 15 days (10 to 42 days) after the onset of symptoms. Due to isolation policies, this visit was often done at the same time than the next study visit. Blood was drawn to study humoral and cellular immunity and a questionnaire focusing on SARS-CoV-2 detection tests done and COVID-19 symptoms was completed at that visit.

### Study exposures and follow-up

The two main exposures of the study were SARS-CoV-2 infection, defined as a positive virus detection test for SARS-CoV-2 (PCR or antigen detection), and SARS-CoV-2 vaccination. Participants were asked about possible or confirmed SARS-CoV-2 infection (i.e., symptoms, virus detection test, date and result) and their SARS-CoV-2 vaccination history (i.e., number of doses, date of vaccination, type of vaccine) since the beginning of the pandemic at V1, and since the last visit (at V2 to V5). Positive SARS-CoV-2 detection tests were therefore captured from the beginning of the pandemic until the earliest among the last visit, withdrawal from the study, or loss of eligibility.

### Study outcome

The primary outcomes were vaccine- and infection-induced immunity. Humoral and cellular immunity (innate and acquired) was explored, using different techniques and antigens.

### Ethics Statement

This study was conducted in accordance with the ethical principles outlined in the Belmont Report and the Declaration of Helsinki. All participants provided written informed consent prior to inclusion in the study. This study was approved by the « Comité d’éthique de la recherche du CHU de Québec – Université Laval » (registration number 2021-5744).

### Confidentiality and data storage

A unique, anonymized identifier was assigned to each participant and used for the data and the blood samples. The samples are stored for up to 10 years, and the data for at least 15 years.

### Participants’ information

More detailed, participant-level information is publicly available on an online platform developed by Maelstrom Research (15). Data on participants’ immune responses to SARS-CoV-2 infection and vaccination are shared through peer-reviewed publications.

### Patient and public involvement statement

No public stakeholders were involved in establishing and designing this cohort.

## Results

### Participant characteristics

Overall, 304 individuals were initially recruited to attend the three first visits (12±2 weeks) from April 20^th^, 2021 to May 9^th^, 2022. The cohort included 149 (49.0%) restaurant/bar workers, 112 (36.8%) grocery store workers, and 43 (14.1%) hardware store workers. With the emergence of Omicron, 198 participants who were still within the recruiting window at the time of ethics approval, were included for two additional visits (12±4 weeks) between March 15^th^, 2022 and October 3^rd^, 2022. Only 13 out of 304 (4.3%) withdrew before V3, and 4 more out of 198 (2.0%) withdrew at V5, resulting in a series of at least 5 blood samples drawn over 48 weeks for most participants.

On average, participants were aged 41.3 years in the overall cohort (Table 2). Specifically, restaurants/bar workers were on average 37.2 years old, grocery store workers 44.2 and hardware store workers 48.2. Participants 60 years of age or older represented 15.5% of the cohort. Female participants represented 57.9% of the cohort. In this cohort, 96.7% self-identified as White, 1.6% as Asian, 1.0% as Latino American, and 0.7% as Black. Levels of education varied: 39.5% reported having a high school diploma or a vocational certificate, 33.2% college education and 22.7% at least a university degree. Participants lived and worked mostly in the Capitale-Nationale administrative region, the remainder being in the Chaudière-Appalaches (Table 3). Most (i.e., 61.8%) lived alone or with one other person, 23.0% lived with children (<18 years), 15.5% with HCWs and 7.6% with teachers or kindergarten workers. These distributions were similar within each occupational group.

**Table 2.** Detailed demographics of study participants at the first visit.

|  | Overall study population |  | Restaurant/bar workers |  | Grocery store workers |  | Hardware store workers |  |
| --- | --- | --- | --- | --- | --- | --- | --- | --- |
|  | Total (N=304) | COVID-19<sup>1</sup> (N=117) | Total (N=149) | COVID-19<sup>1</sup> (N=62) | Total (N=112) | COVID-19<sup>1</sup> (N=42) | Total (N=43) | COVID-19<sup>1</sup> (N=13) |
| **Age (years), Mean±SD** | 41.3 ± 15.9 | 39.6 ± 14.6 | 37.2 ± 14.8 | 36.2 ± 14.3 | 44.2 ± 15.3 | 42.4 ± 14.1 | 48.2 ± 17.3 | 46.5 ± 14.6 |
| Age groups, N (%) |  |  |  |  |  |  |  |  |
| 18-59 | 257 (84.5%) | 106 (90.6%) | 135 (90.6%) | 57 (91.9%) | 93 (83.0%) | 37 (88.1%) | 29 (67.4%) | 12 (92.3%) |
| 60-75 | 47 (15.5%) | 11 (9.4%) | 14 (9.4%) | 5 (8.1%) | 19 (17.0%) | 5 (11.9%) | 14 (32.6%) | 1 (7.7%) |
| **Sex, N (%)** |  |  |  |  |  |  |  |  |
| Female | 176 (57.9%) | 72 (61.5%) | 95 (63.8%) | 39 (62.9%) | 57 (50.9%) | 25 (59.5%) | 24 (55.8%) | 8 (61.5%) |
| Male | 128 (42.1%) | 45 (38.5%) | 54 (36.2%) | 23 (37.1%) | 55 (49.1%) | 17 (40.5%) | 19 (44.2%) | 5 (38.5%) |
| **Race/ethnicity,<sup>2</sup> N (%)** |  |  |  |  |  |  |  |  |
| White | 294 (96.7%) | 114 (97.4%) | 142 (95.3%) | 61 (98.4%) | 109 (97.3%) | 40 (95.2%) | 43 (100.0%) | 13 (100.0%) |
| Asian | 5 (1.6%) | 0 (0.0%) | 4 (2.7%) | 0 (0.0%) | 1 (0.9%) | 0 (0.0%) | 0 (0.0%) | 0 (0.0%) |
| Latino American | 3 (1.0%) | 1 (0.9%) | 3 (2.0%) | 1 (1.6%) | 0 (0.0%) | 0 (0.0%) | 0 (0.0%) | 0 (0.0%) |
| Black | 2 (0.7%) | 2 (1.7%) | 0 (0.0%) | 0 (0.0%) | 2 (1.8%) | 2 (4.8%) | 0 (0.0%) | 0 (0.0%) |
| **Educational attainment, N (%)** |  |  |  |  |  |  |  |  |
| Less than high school | 14 (4.6%) | 3 (2.6%) | 4 (2.7%) | 2 (3.2%) | 9 (8.0%) | 0 (0.0%) | 1 (2.3%) | 1 (7.7%) |
| High school | 79 (26.0%) | 27 (23.1%) | 42 (28.2%) | 14 (22.6%) | 23 (20.5%) | 7 (16.7%) | 14 (32.6%) | 6 (46.2%) |
| Professional | 41 (13.5%) | 13 (11.1%) | 19 (12.8%) | 6 (9.7%) | 16 (14.3%) | 6 (14.3%) | 6 (14.0%) | 1 (7.7%) |
| College | 101 (33.2%) | 45 (38.5%) | 51 (34.2%) | 26 (41.9%) | 35 (31.3%) | 14 (33.3%) | 15 (34.9%) | 5 (38.5%) |
| University baccalaureate | 54 (17.8%) | 22 (18.8%) | 28 (18.8%) | 12 (19.4%) | 21 (18.8%) | 10 (23.8%) | 5 (11.6%) | 0 (0.0%) |
| Graduate studies | 15 (4.9%) | 7 (6.0%) | 5 (3.4%) | 2 (3.2%) | 8 (7.1%) | 5 (11.9%) | 2 (4.7%) | 0 (0.0%) |

1. Subset of participants who reported a positive SARS-CoV-2 test (PCR or rapid antigen) at least once during the study period.
2. Self-reported

**Table 3.**
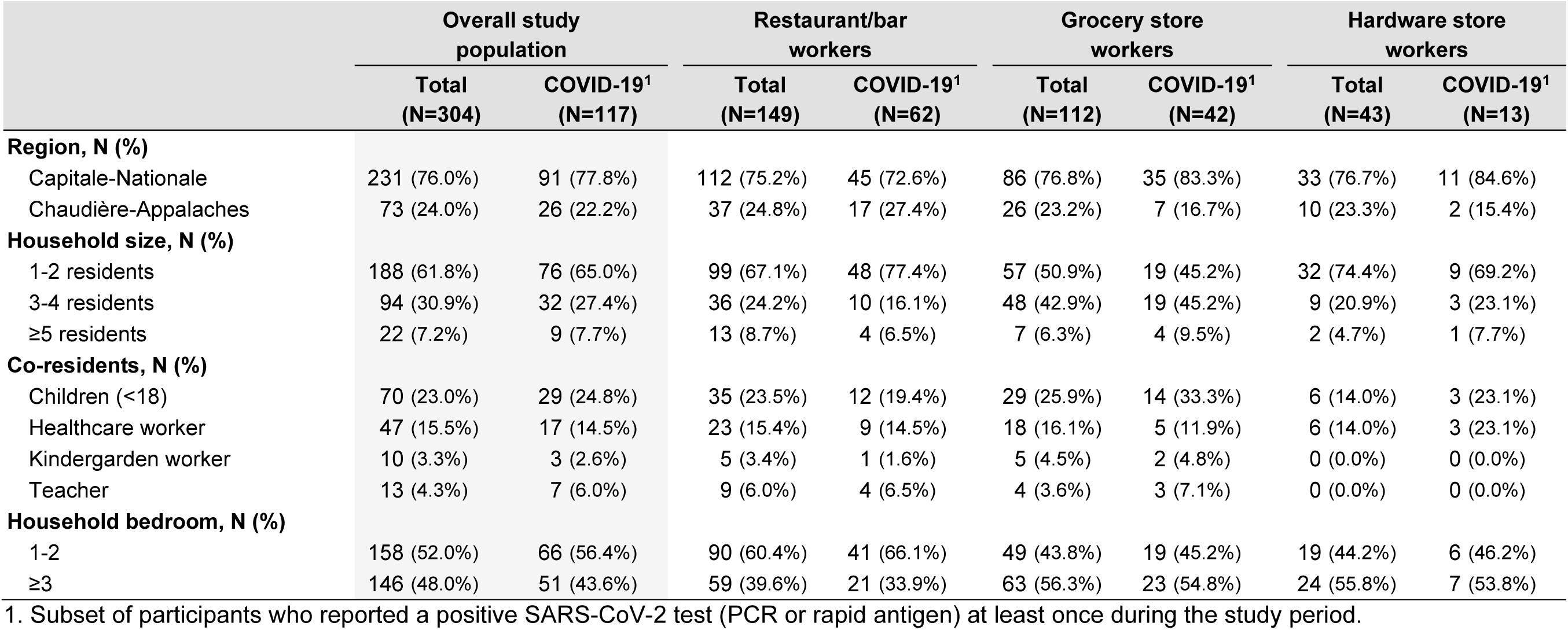
Household characteristics of the participants at the first visit.

According to body mass index (BMI), 41.1% of the participants had a healthy weight (i.e., BMI=18.5 to 24.9 kg/m^2^), 27.0% were overweight (BMI=25.0 to 29.9 kg/m^2^) and 30.6% were obese (BMI ≥30 kg/m^2^) (Table 4). Only 1.3% were underweight (BMI <18.5 kg/m^2^). Cigarette (i.e., tobacco) use was reported by 17.4% of the participants and e-cigarette by 7.9%. About a third (i.e., 31.9%) of participants reported having at least one comorbidity. Hardware store workers had more comorbidities (53.8%), probably because they were slightly older. Overall, 17.1% reported usually receiving the annual influenza vaccine and 13.8% in the year prior to the first visit.

**Table 4.**
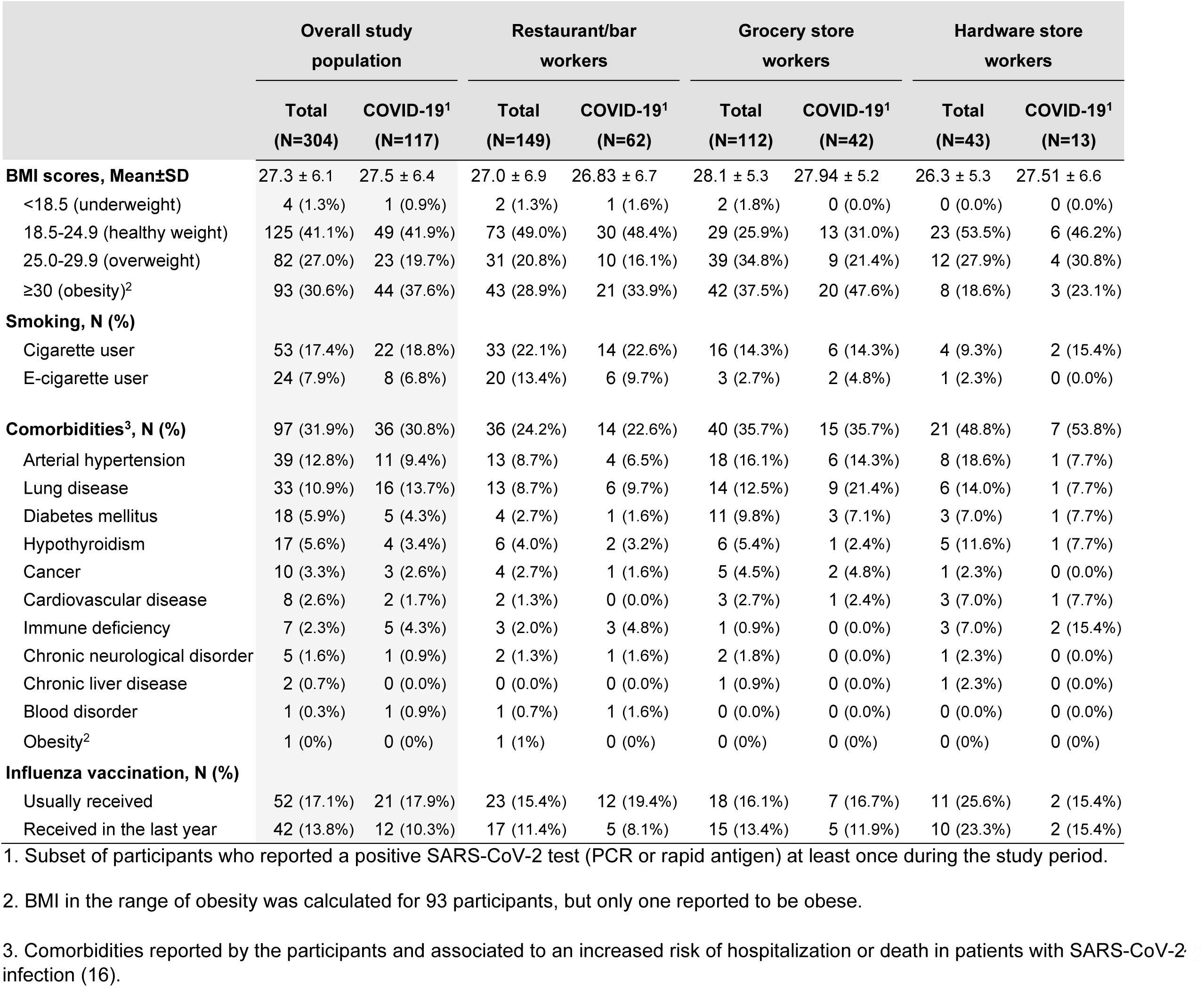
Clinical characteristics of study participants at the first visit.

Approximately half (i.e., 46.4%) of the participants reported working on average more than 30 hours per week (Table 5). Most (88.8%) had attended at least one gathering of 10 or more persons during the study period, and 40.8% attended more than 10 such gatherings. The predominant mode of transportation was by car (87.5%), followed by bus (12.8%) and walking (9.2%). Traveling outside the province of Québec was reported by 47.0% of the participants, with 25.7% travelling within Canada, 14.8% to the United States, and 27.6% elsewhere. The distribution of participants in each occupational group was similar for the workplace region, mode of transportation and travelling, but differed for the weekly hours worked and the number of gatherings attended.

**Table 5.** Occupational and behavioral characteristic of study participants.

|  | Overall study population |  | Restaurant/bar workers |  | Grocery store workers |  | Hardware store workers |  |
| --- | --- | --- | --- | --- | --- | --- | --- | --- |
|  | Total (N=304) | COVID-19<sup>1</sup> (N=117) | Total (N=149) | COVID-19<sup>1</sup> (N=62) | Total (N=112) | COVID-19<sup>1</sup> (N=42) | Total (N=43) | COVID-19<sup>1</sup> (N=13) |
| **Workplace region,<sup>2</sup> N (%)** |  |  |  |  |  |  |  |  |
| Capitale-Nationale | 240 (78.9%) | 96 (82.1%) | 123 (82.6%) | 49 (79.0%) | 85 (75.9%) | 36 (85.7%) | 32 (74.4%) | 11 (84.6%) |
| Chaudière-Appalaches | 64 (21.1%) | 19 (16.2%) | 26 (17.4%) | 12 (19.4%) | 27 (24.1%) | 6 (14.3%) | 11 (25.6%) | 1 (7.7%) |
| **Weekly hours worked,<sup>3</sup> Mean±SD** | 27.9 ± 11.5 | 28.9 ± 11.4 | 24.7 ± 11.0 | 24.5 ± 11.0 | 32.6 ± 10.4 | 34.2 ± 9.8 | 26.6 ± 11.9 | 32.3 ± 10.4 |
| Participants working |  |  |  |  |  |  |  |  |
| Full time (≥30) | 141 (46.4%) | 64 (54.7%) | 49 (32.9%) | 23 (37.1%) | 73 (65.2%) | 32 (76.2%) | 19 (44.2%) | 9 (69.2%) |
| Part time (<30) | 163 (53.6%) | 53 (45.3%) | 100 (67.1%) | 39 (62.9%) | 39 (34.8%) | 10 (23.8%) | 24 (55.8%) | 4 (30.8%) |
| **Gathering of 10+ persons,<sup>3</sup> Mean±SD** |  |  |  |  |  |  |  |  |
| Per group, N (%) |  |  |  |  |  |  |  |  |
| None | 34 (11.2%) | 6 (5.1%) | 16 (10.7%) | 2 (3.2%) | 15 (13.4%) | 4 (9.5%) | 3 (7.0%) | 0 (0.0%) |
| 1 to 10 gatherings | 146 (48.0%) | 47 (40.2%) | 57 (38.3%) | 16 (25.8%) | 62 (55.4%) | 22 (52.4%) | 27 (62.8%) | 9 (69.2%) |
| 11 to 50 gatherings | 101 (33.2%) | 51 (43.6%) | 58 (38.9%) | 33 (53.2%) | 32 (28.6%) | 14 (33.3%) | 11 (25.6%) | 4 (30.8%) |
| >50 gatherings | 23 (7.6%) | 13 (11.1%) | 18 (12.1%) | 11 (17.7%) | 3 (2.7%) | 2 (4.8%) | 2 (4.7%) | 0 (0.0%) |
| **Transportation,<sup>2</sup> N (%)** |  |  |  |  |  |  |  |  |
| Car | 266 (87.5%) | 103 (88.0%) | 127 (85.2%) | 54 (87.1%) | 98 (87.5%) | 37 (88.1%) | 41 (95.3%) | 12 (92.3%) |
| Bus | 39 (12.8%) | 16 (13.7%) | 23 (15.4%) | 9 (14.5%) | 10 (8.9%) | 5 (11.9%) | 6 (14.0%) | 2 (15.4%) |
| Walking | 28 (9.2%) | 11 (9.4%) | 13 (8.7%) | 6 (9.7%) | 14 (12.5%) | 5 (11.9%) | 1 (2.3%) | 0 (0.0%) |
| Bicycle | 13 (4.3%) | 6 (5.1%) | 7 (4.7%) | 3 (4.8%) | 4 (3.6%) | 2 (4.8%) | 2 (4.7%) | 1 (7.7%) |
| Carpooling | 2 (0.7%) | 2 (1.7%) | 1 (0.7%) | 1 (1.6%) | 1 (0.9%) | 1 (2.4%) | 0 (0.0%) | 0 (0.0%) |
| **Travel,<sup>3</sup> N (%)** |  |  |  |  |  |  |  |  |
| Any destination | 143 (47.0%) | 70 (59.8%) | 79 (53.0%) | 40 (64.5%) | 48 (42.9%) | 25 (59.5%) | 16 (37.2%) | 5 (38.5%) |
| In Canada | 78 (25.7%) | 39 (33.3%) | 45 (30.2%) | 24 (38.7%) | 23 (20.5%) | 13 (31.0%) | 10 (23.3%) | 2 (15.4%) |
| To USA | 45 (14.8%) | 20 (17.1%) | 25 (16.8%) | 11 (17.7%) | 16 (14.3%) | 8 (19.0%) | 4 (9.3%) | 1 (7.7%) |
| Other destination<sup>4</sup> | 84 (27.6%) | 45 (38.5%) | 47 (31.5%) | 25 (40.3%) | 28 (25.0%) | 16 (38.1%) | 9 (20.9%) | 4 (30.8%) |

1. Subset of participants who reported a positive SARS-CoV-2 test (PCR or rapid antigen) at least once during the study period.
2. At the time of the first visit.
3. During the entire study period.
4. Includes travel to Cuba, Ireland, Great-Britain, Luxembourg, Dominican Republic, South Africa, Bahamas, Morocco, Guadeloupe, Panama, Costa Rica, Greece.

Overall, 98.4% of the participants reported wearing a mask at work, indicating excellent adherence to this measure (Table 6). Other measures at work, such as handwashing (98.4%), social distancing (70.1%) and the use of Plexiglas dividers (77.3%) were also frequent. The use of gloves (6.9%) and face-shields (10.5%), which were not extensively promoted by the public health authorities, were less frequent. Outside work, all participants reported wearing a mask in public (100.0%); most avoided usual salutations (85.2%), practiced social distancing (84.2%) and avoided contact with vulnerable persons (83.6%) and crowded places (76.6%). Most participants reported washing their hands when dirty (96.7%), after using the restroom (97.7%), when arriving at (92.1%) and leaving the workplace (71.7%), before eating (87.8%) and after handling trash (79.6%). In general, adherence to these measures was consistently lower among restaurant and bar workers, possibly because of the nature of their work or their younger age (on average).

**Table 6.** Protective measures taken at work and elsewhere by study participants recorded at first visit.

|  | Overall study population |  | Restaurant/bar workers |  | Grocery store workers |  | Hardware store workers |  |
| --- | --- | --- | --- | --- | --- | --- | --- | --- |
|  | Total (N=304) | COVID-19<sup>1</sup> (N=117) | Total (N=149) | COVID-19<sup>1</sup> (N=62) | Total (N=112) | COVID-19<sup>1</sup> (N=42) | Total (N=43) | COVID-19<sup>1</sup> (N=13) |
| **Protection measures at work,<sup>2</sup> N (%)** |  |  |  |  |  |  |  |  |
| Mask | 299 (98.4%) | 116 (99.1%) | 147 (98.7%) | 61 (98.4%) | 111 (99.1%) | 42 (100.0%) | 41 (95.3%) | 13 (100.0%) |
| Handwashing | 299 (98.4%) | 114 (97.4%) | 145 (97.3%) | 59 (95.2%) | 111 (99.1%) | 42 (100.0%) | 43 (100.0%) | 13 (100.0%) |
| Plexiglas | 235 (77.3%) | 88 (75.2%) | 96 (64.4%) | 39 (62.9%) | 98 (87.5%) | 37 (88.1%) | 41 (95.3%) | 12 (92.3%) |
| Social distancing | 213 (70.1%) | 86 (73.5%) | 111 (74.5%) | 48 (77.4%) | 74 (66.1%) | 29 (69.0%) | 28 (65.1%) | 9 (69.2%) |
| Protective glasses | 77 (25.3%) | 12 (10.3%) | 30 (20.1%) | 4 (6.5%) | 35 (31.3%) | 5 (11.9%) | 12 (27.9%) | 3 (23.1%) |
| Faceshield | 32 (10.5%) | 4 (3.4%) | 16 (10.7%) | 1 (1.6%) | 13 (11.6%) | 2 (4.8%) | 3 (7.0%) | 1 (7.7%) |
| Gloves | 21 (6.9%) | 3 (2.6%) | 10 (6.7%) | 0 (0.0%) | 9 (8.0%) | 3 (7.1%) | 2 (4.7%) | 0 (0.0%) |
| Face Cover | 7 (2.3%) | 1 (0.9%) | 3 (2.0%) | 0 (0.0%) | 4 (3.6%) | 1 (2.4%) | 0 - | 0 (0.0%) |
| Other<sup>3</sup> | 135 (44.4%) | 59 (50.4%) | 85 (57.0%) | 41 (66.1%) | 43 (38.4%) | 16 (38.1%) | 7 (16.3%) | 2 (15.4%) |
| **Behavioral protection measures,<sup>2</sup> N (%)** |  |  |  |  |  |  |  |  |
| Mask wearing in public spaces | 304 (100.0%) | 117 (100.0%) | 149 (100.0%) | 62 (100.0%) | 112 (100.0%) | 42 (100.0%) | 43 (100.0%) | 12 (92.3%) |
| Avoid usual salutations | 259 (85.2%) | 99 (84.6%) | 119 (79.9%) | 48 (77.4%) | 103 (92.0%) | 39 (92.9%) | 37 (86.0%) | 12 (92.3%) |
| Social distancing | 256 (84.2%) | 93 (79.5%) | 115 (77.2%) | 41 (66.1%) | 102 (91.1%) | 40 (95.2%) | 39 (90.7%) | 12 (92.3%) |
| Avoid contacts with vulnerable persons | 254 (83.6%) | 93 (79.5%) | 114 (76.5%) | 42 (67.7%) | 100 (89.3%) | 38 (90.5%) | 40 (93.0%) | 13 (100.0%) |
| Avoid crowded places | 233 (76.6%) | 81 (69.2%) | 102 (68.5%) | 35 (56.5%) | 94 (83.9%) | 34 (81.0%) | 37 (86.0%) | 12 (92.3%) |
| Quarantine if exposed to COVID-19 | 126 (41.4%) | 70 (59.8%) | 65 (43.6%) | 36 (58.1%) | 46 (41.1%) | 25 (59.5%) | 15 (34.9%) | 9 (69.2%) |
| Pre-emptive isolation | 36 (11.8%) | 23 (19.7%) | 21 (14.1%) | 13 (21.0%) | 12 (10.7%) | 9 (21.4%) | 3 (7.0%) | 1 (7.7%) |
| **Handwashing habits,<sup>2</sup> N (%)** |  |  |  |  |  |  |  |  |
| After using the restroom | 297 (97.7%) | 115 (98.3%) | 147 (98.7%) | 60 (96.8%) | 109 (97.3%) | 42 (100.0%) | 41 (95.3%) | 13 (100.0%) |
| When dirty | 294 (96.7%) | 113 (96.6%) | 140 (94.0%) | 58 (93.5%) | 112 (100.0%) | 42 (100.0%) | 42 (97.7%) | 13 (100.0%) |
| When entering workspace | 280 (92.1%) | 106 (90.6%) | 136 (91.3%) | 55 (88.7%) | 107 (95.5%) | 37 (88.1%) | 37 (86.0%) | 13 (100.0%) |
| Before eating | 267 (87.8%) | 104 (88.9%) | 124 (83.2%) | 51 (82.3%) | 105 (93.8%) | 41 (97.6%) | 38 (88.4%) | 12 (92.3%) |
| Before & after handling food | 246 (80.9%) | 96 (82.1%) | 127 (85.2%) | 55 (88.7%) | 93 (83.0%) | 32 (76.2%) | 26 (60.5%) | 9 (69.2%) |
| After handling trash | 242 (79.6%) | 97 (82.9%) | 119 (79.9%) | 51 (82.3%) | 95 (84.8%) | 38 (90.5%) | 28 (65.1%) | 8 (61.5%) |
| When exiting workspace | 218 (71.7%) | 81 (69.2%) | 104 (69.8%) | 41 (66.1%) | 87 (77.7%) | 32 (76.2%) | 27 (62.8%) | 8 (61.5%) |
| Other<sup>3</sup> | 50 (16.4%) | 11 (9.4%) | 29 (19.5%) | 4 (6.5%) | 12 (10.7%) | 2 (4.8%) | 9 (20.9%) | 5 (38.5%) |

1. Subset of participants who reported a positive SARS-CoV-2 test (PCR or rapid antigen) at least once during the study period.
2. Includes customer registry, QR code, customer limit in store, thorough cleaning of workplace, worker temperature surveillance.
3. Includes after touching the cash register, handling money, in between clients

### SARS-CoV-2 infection and related symptoms

A total of 117 participants (38.5%) reported at least one positive test (PCR or rapid antigen) indicating a first infection (Table 7). Eight participants experienced a second positive test (occurring more than 90 days after a previous positive result) during the study. Among these 117 participants with a first infection, 94.9% reported at least one symptom at the time of testing (Table 8). Each individual symptom was experienced by ≥47.9% of participants, except for diarrhea (13.7%) and loss of smell or taste (22.2%), a pattern consistent with prior studies (17–19). These distributions were similar within each occupational group. A sub-group of 29 participants had symptoms that made them to suspect that they had COVID-19, but PCR or antigen detection test were negative or not done. Routine PCR tests conducted on asymptomatic participants during visits 4 and 5 returned positive on seven occasions. Of these, four participants had never tested positive, while three had tested positive more than 90 days earlier.

**Table 7.** Number of participants with a first SARS-CoV-2 positive test.

|  | Overall cohort | Bar/<br>Restaurant | Grocery store | Hardware store |
| --- | --- | --- | --- | --- |
| **Participant reporting** | **117** | **62** | **42** | **13** |
| PCR | 41 | 25 | 14 | 2 |
| Antigen detection | 76 | 37 | 28 | 11 |

**Table 8.**
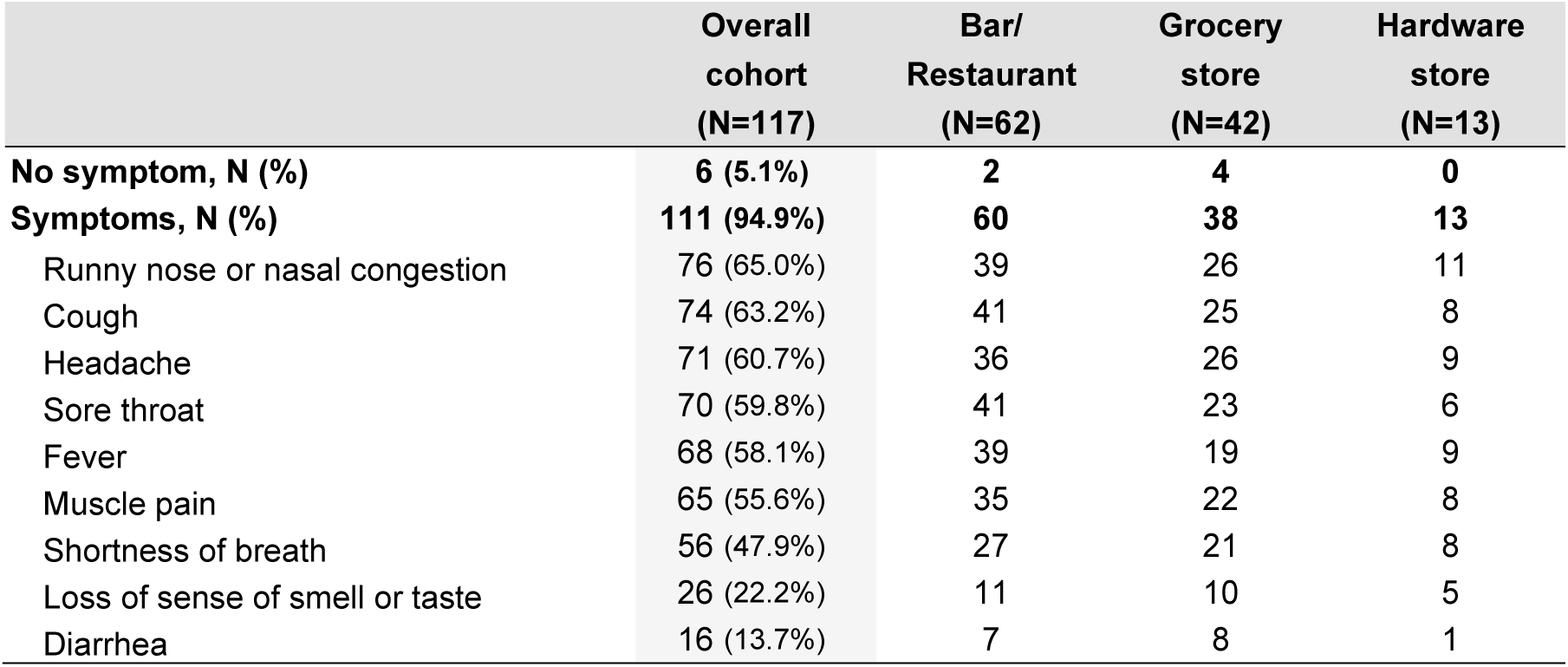
COVID-19 symptoms at the first SARS-CoV-2 positive test.

|  | Overall cohort<br>(N=117) | Bar/<br>Restaurant<br>(N=62) | Grocery<br>store<br>(N=42) | Hardware<br>store<br>(N=13) |
| --- | --- | --- | --- | --- |
| **No symptom, N (%)** | **6 (5.1%)** | **2** | **4** | **0** |
| **Symptoms, N (%)** | **111 (94.9%)** | **60** | **38** | **13** |
| Runny nose or nasal congestion | 76 (65.0%) | 39 | 26 | 11 |
| Cough | 74 (63.2%) | 41 | 25 | 8 |
| Headache | 71 (60.7%) | 36 | 26 | 9 |
| Sore throat | 70 (59.8%) | 41 | 23 | 6 |
| Fever | 68 (58.1%) | 39 | 19 | 9 |
| Muscle pain | 65 (55.6%) | 35 | 22 | 8 |
| Shortness of breath | 56 (47.9%) | 27 | 21 | 8 |
| Loss of sense of smell or taste | 26 (22.2%) | 11 | 10 | 5 |
| Diarrhea | 16 (13.7%) | 7 | 8 | 1 |

### Vaccination for SARS-CoV-2

The participants were vaccinated according to recommendations with the vaccines approved by Canadian health authorities: monovalent Comirnaty (Pfizer-BioNTech), Spikevax (Moderna) and Vaxzevria (AstraZeneca), which each required two doses to complete the primary series. By the end of the study (i.e., last visit between May 10^th^, 2022 and October 3^rd^, 2022), 95.9% of the participants (Fig 2) were fully vaccinated (at least 2 doses) and nearly 70% had received at least one booster dose. Hardware store workers were the most highly vaccinated occupational group, 98.6% of them having received two doses early in the study. In participants having completed the primaries series of vaccine, all infections occurred after the 4^th^ wave, when the Omicron variant was predominant.

**Figure 2.**
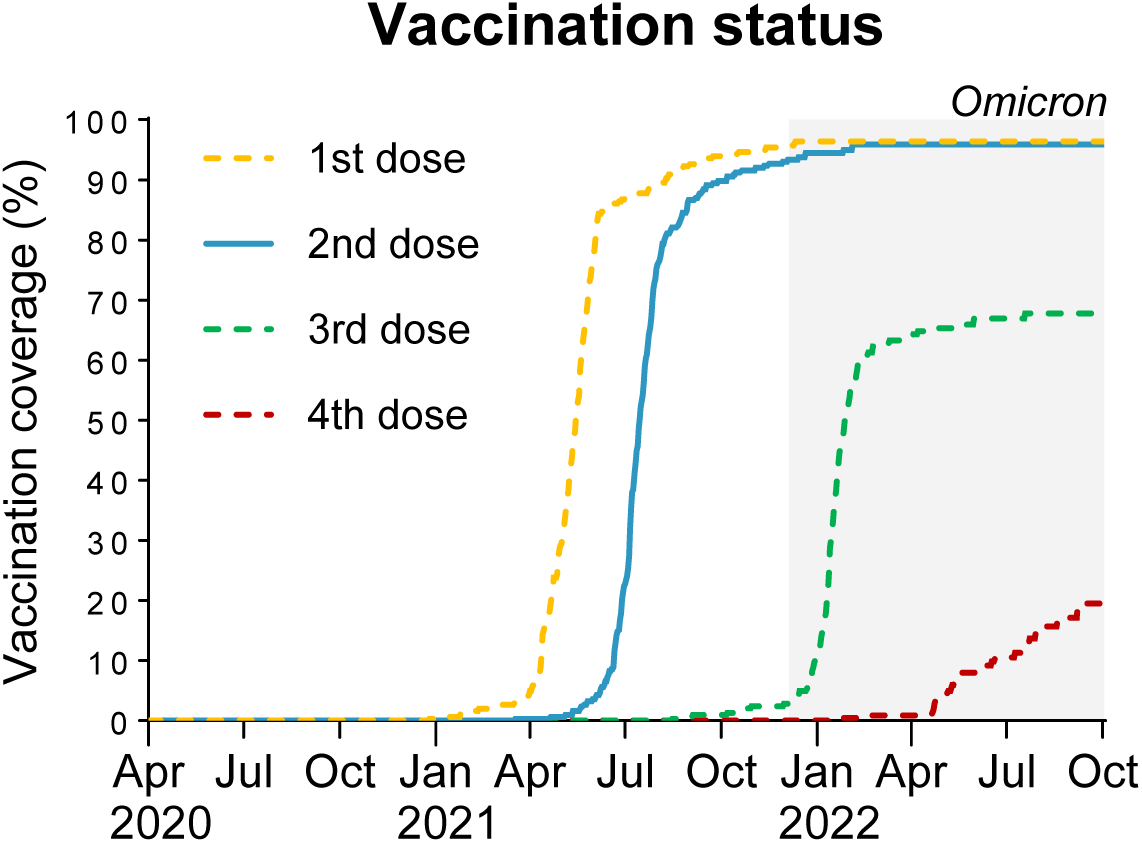
Evolution of the vaccination coverage stratified by the number of doses. Data were generated using Kaplan-Meier analysis to account for variation in follow-up duration. Shaded area represents Omicron predominance.

### Blood sample bank to study infection-induced, vaccine-induced, and hybrid immunity

Overall, 1299 blood samples were collected, including 304 (23.4%) at V1, 297 (22.9%) at V2, 291 (22.4%) at V3, 198 (15.2%) at V4, 194 (15.0%) at V5, and 15 (1.2%) at additional visits (i.e., VCoV). In total, 69.2% of the blood samples were drawn from vaccinated participants with no known history of SARS-CoV-2 infection, 23.1% from vaccinated and previously infected participants, 6.9% from unvaccinated participants with no known history of SARS-CoV-2 infection, and 11 (0.8%) from previously infected and unvaccinated participants.

### Published studies from this cohort

Five studies using the data collected by the cohort has been recently published:

- The study of infection rate and risk factors for SARS-CoV-2 infection among retail workers, focusing on the impact of emerging Omicron variants and workplace protective measures (20).
- The impact of SARS-CoV-2 vaccination and seasonal variations on the activation of innate immune cells by the viral analog R848, which targets TLR7/8 (21).
- The neutralization activity of SARS-CoV-2 antibodies was measured against various strains in 280 vaccinated participants (22).
- The investigation of the humoral response to vaccination and SARS-CoV-2 infection to identify factors influencing the serological response using newly developed ELISA assays (23).
- The validation of a plasmonic sensor for clinical use aimed to assess its ability to measure SARS-CoV-2 “pseudo-neutralization” demonstrating its correlation with live virus assays and epidemiological data (24).

## Conclusions, strengths and limitations

We set up a cohort of 304 participants to conduct a longitudinal study of SARS-CoV-2 infection rate and immunity among food and retail workers who lived and worked within the Québec City metropolitan region. The participants provided information on a wide range of demographic, socioeconomic, behavioral, occupational and clinical variables. Information on COVID-19 symptoms (when applicable), as well as SARS-CoV-2 vaccination status and any SARS-CoV-2 positive diagnostic test (i.e., PCR or rapid antigen) from the beginning of the pandemic until the last visit was also collected. The study was initiated at a time when all public health measures were implemented and covered seven waves of COVID-19 infection, including those dominated by the Ancestral strain, as well as Alpha, Delta, and Omicron variants, thus capturing a relatively large number of epidemiological periods and infections. In addition, blood samples were collected at each scheduled visit regardless of participants’ infection or vaccination history, thus enabling the study of infection-induced, vaccine-induced and hybrid immunity in this extensively characterized cohort. Moreover, very few participants withdrew from the study before the end of the initial (i.e., V1-V3) and extension phases (i.e., V4-V5), resulting in complete series of at least 5 samples for most participants.

A total of 117 first (ever) COVID-19 infections, as determined by reported positive PCR or antigen detection test; this represents 38% of the participants. Most of these infections were symptomatic (94.9%) and occurred between December 5^th^, 2021 and October 3^rd^, 2022, consistent with the emergence of the highly contagious Omicron variants. In the present study, vaccine coverage was high: by the time Omicron had emerged, nearly 95% of the participants had already received two vaccine doses (primary series). This high rate of vaccination may be because retail workers considered themselves at higher risk of SARS-CoV-2 exposure than the general population and were thus more willing to get vaccinated and reduce their risk of infection.

Some limitations must be considered when interpreting our results. Per study design, none of the participants had previously experienced a severe COVID-19 illness that required hospitalization. Therefore, the cohort may not be used to study the immune response that leads to severe health outcomes but is appropriate to study the immune response to mild COVID-19 illness as experienced by most patients. Another limitation is that the cohort may have been subject to a sampling bias as people willing to participate in a scientific study may be less apprehensive about being vaccinated. Hence, the study participants may not be representative of the overall population of workers in these sectors. This is suggested by the 5% to 7% higher vaccination coverage for the second dose as of fall 2021 compared to the general population of the province of Québec (25).

The low proportion of racial minorities (i.e., 3.3%) is consistent with the size of the visible minority population in the Québec City metropolitan area (i.e., 4.9% according to census), but limits the use of this cohort to study racial determinants of immunity to SARS-CoV-2. In addition, participant responses on previous COVID-19 may have been affected by a memory bias, particularly for those whose last infection occurred months before V1. Moreover, few samples were drawn from unvaccinated and previously infected participants, so that the cohort may be of limited use to study immunity induced by infection alone. The high vaccination coverage also made it impossible to assess the impact of vaccination on the risk of infection, since, at any given time, most participants were vaccinated. Lastly, our study may have underestimated the incidence of SARS-CoV-2 infection, as most cases occurred during the Omicron wave when access to PCR testing was limited, antigen tests were less sensitive, and infections were self-reported rather than identified through laboratory surveillance. The serology data of these samples will shed light on this question.

## Data Availability

All data produced in the present study are available upon reasonable request to the authors
All data produced are available online at the COVID-19 Immunity Task Force (CITF) Data Portal

https://portal.citf.mcgill.ca/

## Acknowledgements

The authors thank the participants and all the staff involved in planning and preparation of this study. David Simonyan helped with statistical analysis. A special thank you to our partners: CSN Federation of Commerce and AQMAT.

## Author contributions

**Conceptualization:** Joelle N. Pelletier, Caroline Gilbert, Jean-François Masson, Mariana Baz, Denis Boudreau, Sylvie Trottier

**Data Curation:** Mathieu Thériault, Kim Santerre, SylvieTrottier.

**Formal Analysis:** Mathieu Thériault, Nicholas Brousseau, Samuel Rochette, Sylvie Trottier.

**Funding Acquisition:** Denis Boudreau, Sylvie Trottier

**Investigation:** Mathieu Thériault, Kim Santerre, Samuel Rochette, Sylvie Trottier.

**Methodology:** Kim Santerre, Samuel Rochette, Sylvie Trottier

**Project Administration:** Denis Boudreau, Sylvie Trottier.

**Resources:** Denis Boudreau, Sylvie Trottier.

**Software:** Mathieu Thériault.

**Supervision**: Denis Boudreau, SylvieTrottier.

**Validation:** Mathieu Thériault, Kim Santerre, Nicholas Brousseau, Samuel Rochette, Rabeea F. Omar, Joelle N. Pelletier, Caroline Gilbert, Jean-François Masson, Mariana Baz, Denis Boudreau, Sylvie Trottier.

**Visualization:** Mathieu Thériault, Kim Santerre, Samuel Rochette, Sylvie Trottier

**Writing:** Mathieu Thériault, Kim Santerre, Samuel Rochette, Sylvie Trottier.

**Writing – Review & Editing :** Mathieu Thériault, Kim Santerre, Nicholas Brousseau, Samuel Rochette, Rabeea F. Omar, Joelle N. Pelletier, Caroline Gilbert, Jean-François Masson, Mariana Baz, Denis Boudreau, Sylvie Trottier.

## REFERENCES

1. Pathela P, Crawley A, Weiss D, Maldin B, Cornell J, Purdin J, et al. Seroprevalence of Severe Acute Respiratory Syndrome Coronavirus 2 Following the Largest Initial Epidemic Wave in the United States: Findings From New York City, 13 May to 21 July 2020. J Infect Dis. 2021;224(2):196–206.

2. Feehan AK, Velasco C, Fort D, Burton JH, Price-Haywood EG, Katzmarzyk PT, et al. Racial and Workplace Disparities in Seroprevalence of SARS-CoV-2, Baton Rouge, Louisiana, USA. Emerg Infect Dis. 2021;27(1).

3. Sim MR. The COVID-19 pandemic: major risks to healthcare and other workers on the front line. Occup Environ Med. 2020;77(5):281–2.

4. Gomez-Ochoa SA, Franco OH, Rojas LZ, Raguindin PF, Roa-Diaz ZM, Wyssmann BM, et al. COVID-19 in Health-Care Workers: A Living Systematic Review and Meta-Analysis of Prevalence, Risk Factors, Clinical Characteristics, and Outcomes. Am J Epidemiol. 2021;190(1):161–75.

5. Gholami M, Fawad I, Shadan S, Rowaiee R, Ghanem H, Hassan Khamis A, et al. COVID-19 and healthcare workers: A systematic review and meta-analysis. Int J Infect Dis. 2021;104:335–46.

6. Boucher E, Cao C, D’Mello S, Duarte N, Donnici C, Duarte N, et al. Occupation and SARS-CoV-2 seroprevalence studies: a systematic review. BMJ Open. 2023;13(2):e063771.

7. Vachon MS, Demmer RT, Yendell S, Draeger KJ, Beebe TJ, Hedberg CW. SARS-CoV-2 Seroprevalence Survey in Grocery Store Workers-Minnesota, 2020-2021. Int J Environ Res Public Health. 2022;19(6).

8. Parks CA, Nugent NB, Fleischhacker SE, Yaroch AL. Food System Workers are the Unexpected but Under Protected COVID Heroes. J Nutr. 2020;150(8):2006–8.

9. Stringhini S, Zaballa ME, Pullen N, de Mestral C, Perez-Saez J, Dumont R, et al. Large variation in anti-SARS-CoV-2 antibody prevalence among essential workers in Geneva, Switzerland. Nat Commun. 2021;12(1):3455.

10. de Gier B, de Oliveira Bressane Lima P, van Gaalen RD, de Boer PT, Alblas J, Ruijten M, et al. Occupation- and age-associated risk of SARS-CoV-2 test positivity, the Netherlands, June to October 2020. Euro Surveill. 2020;25(50).

11. Furuse Y, Sando E, Tsuchiya N, Miyahara R, Yasuda I, Ko YK, et al. Clusters of Coronavirus Disease in Communities, Japan, January-April 2020. Emerg Infect Dis. 2020;26(9).

12. Decarie Y, Michaud PC. Counting the Dead: COVID-19 and Mortality in Quebec and British Columbia During the First Wave. Can Stud Popul. 2021;48(2-3):139–64.

13. Shim E. Regional Variability in COVID-19 Case Fatality Rate in Canada, February-December 2020. Int J Environ Res Public Health. 2021;18(4).

14. COVID-19 Immunity Task Force standardized core survey data elements [Available from: https://www.covid19immunitytaskforce.ca/covid-19-immunity-task-force-releases-standardized-core-survey-data-elements/.

15. CISACOV investigators. Cellular Immunity and Seroprevalence of Antibodies against SARS-CoV-2: Characterisation of Three Populations of Food Workers [Internet]. 2021-2022. Available from: https://www.maelstrom-research.org/study/cisacov.

16. Simard M, Montigny Cd, Jean S, Fortin É, Blais C, Théberge I, et al. Impact des comorbidités sur les risques de décès et d’hospitalisation chez les cas confirmés de la COVID-19 durant les premiers mois de la pandémie au Québec. Institut national de santé publique du Québec; 2020 14 décembre 2020. Report No.: 978-2-550-88147-6.

17. Zhu J, Ji P, Pang J, Zhong Z, Li H, He C, et al. Clinical characteristics of 3062 COVID-19 patients: A meta-analysis. J Med Virol. 2020;92(10):1902–14.

18. Burke RM, Killerby ME, Newton S, Ashworth CE, Berns AL, Brennan S, et al. Symptom Profiles of a Convenience Sample of Patients with COVID-19 - United States, January-April 2020. MMWR Morb Mortal Wkly Rep. 2020;69(28):904–8.

19. Government C. COVID-19 signs, symptoms and severity of disease: A clinician guide. [Available from: https://www.canada.ca/en/public-health/services/diseases/2019-novel-coronavirus-infection/guidance-documents/signs-symptoms-severity.html.

20. Santerre K, Thériault M, Brousseau N, Langlois M-A, Arnold C, Pelletier JN, et al. Infection Rate and Risk Factors of SARS-CoV-2 Infection in Retail Workers at the Onset of the COVID-19 Pandemic, Quebec, Canada. Infectious Disease Reports [Internet]. 2024; 16(6):[1240–53 pp.].

21. Jarras H, Blais I, Goyer B, Bazie WW, Rabezanahary H, Theriault M, et al. Impact of SARS-CoV-2 vaccination and of seasonal variations on the innate immune inflammatory response. Front Immunol. 2024;15:1513717.

22. Rabezanahary H, Gilbert C, Santerre K, Scarrone M, Gilbert M, Theriault M, et al. Live virus neutralizing antibodies against pre and post Omicron strains in food and retail workers in Quebec, Canada. Heliyon. 2024;10(10):e31026.

23. Djaileb A, Parker MF, Lavallee E, Stuible M, Durocher Y, Theriault M, et al. Longitudinal determination of seroprevalence and immune response to SARS-CoV-2 in a population of food and retail workers through decentralized testing and transformation of ELISA datasets. PLoS One. 2024;19(12):e0314499.

24. Coutu J, Ricard P, Djaïleb A, Lavallée É, Rabezanahary H, Stuible M, et al. Large-scale validation of a plasmonic sensor for SARS-CoV-2 pseudo-neutralization with a cohort of food and retail workers. Sensors & Diagnostics. 2024;3(5):850–62.

25. Canada Go. COVID-19 vaccination in Canada: Government of Canada; 2023 [updated 2023-03-03. Available from: https://health-infobase.canada.ca/covid-19/vaccination-coverage/.

